# Genetic Ethnicity and Hypertension Epistatic Interaction Underlying Racial Disparities in US Multiple Myeloma Susceptibility

**DOI:** 10.1101/2024.06.01.24308328

**Authors:** Emmanuel LP Dumont, Luke Han, Srisundesh Kodali, Ariel Aptekmann, Lisa Carter-Bawa, Rena Feinman, Benjamin Tycko, David S. Siegel, Andre Goy, Peter Kaplan, Catherine Do

## Abstract

**Background:** Multiple myeloma (MM), a malignant plasma cell disorder, exhibits pronounced racial disparities in incidence and patient outcomes. The Centers for Disease Control and Prevention (CDC) reports that MM is twice as common in Black Americans as in White Americans. Understanding these racial disparities is paramount to addressing potential healthcare biases and developing targeted interventions to ensure equitable patient care and outcomes.

**Methods:** Using the ‘All of Us’ database from the National Institute of Health, we performed a retrospective study on 413,457 participants. Of these, 1,430 were diagnosed with MM. We examined the factors contributing to racial disparities in MM risk using multivariable statistical analysis, including interaction effects.

**Results:** To comprehensively account for the multidimensional aspects of self-reported race followed by the CDC, we incorporated genetic ethnicity, demographics (age, gender), body mass index, social determinants of health (zipcode’s deprivation index, and health insurance status), and common pre-existing comorbidities (hypertension, diabetes, congestive heart failure - CHF, and chronic obstructive pulmonary disease) into our analysis. Our findings reveal that the racial disparities in health outcomes between non-Hispanic Black and non-Hispanic White individuals, as reported by the CDC, are driven by a synergistic epistatic interaction between having African as a predominant genetic ethnicity and being diagnosed or treated for hypertension (OR: 2.92, 95% CI: 1.54 to 5.57, P = 0.001). This interaction is also true for individuals whose primary genetic ancestry is Ad Mixed American (OR: 2.31, 95% CI: 1.02 to 5.2, P = 0.044). The other variables significantly associated with MM risk are having a predominant genetic ancestry of Ad Mixed American (OR: 0.41, 95% CI: 0.2 to 0.85, P = 0.017), the lack of health insurance (OR: 0.67, 95% CI: 0.48 to 0.93, P = 0.017), zipcode’s deprivation index being above the US median (OR: 1.26, 95% CI: 1.04 to 1.53, P = 0.018), being diagnosed with CHF before MM (OR: 2.06, 95% CI: 1.56 to 2.72, P < 1.e-3), being male (OR: 1.22, 95% CI: 1.02 to 1.46, P = 0.031), and being over the age of 65 (OR: 1.65, 95% CI: 1.36 to 2, P < 1.e-3).

**Conclusion:** These findings reveal a previously unknown epistatic interaction between an individual’s predominant genetic ancestry and hypertension, responsible for the CDC-reported higher risk of the African-American population for MM. In other words, hypertension serves as a surrogate marker for a genetic predisposition in individuals with a predominant African genetic ancestry. This insight could improve the screening and identification of minority individuals at risk for MM.

## INTRODUCTION

Multiple myeloma (MM) is a hematologic malignancy characterized by the accumulation of abnormal plasma cells in the bone marrow, leading to various complications such as marrow failure, skeletal issues, hypercalcemia, and renal dysfunction. MM remains a terminal illness, with around 35,000 new cases diagnosed annually in the United States, making it the second most common blood cancer.^1^ While the 5-year relative survival rate has improved to over 50% in recent years, racial disparities persist in both incidence and patient outcomes.^2–4^

In our article, ‘Whites’ refers to non-Hispanic Whites, ‘Blacks’ refers to non-Hispanic Blacks, and ‘Hispanics’ refers to participants who did not self-identify a race but identified their ethnicity as Hispanic. We use the terms ‘men’ and ‘women’ to refer to participants’ sex assigned at birth, and we use ‘sex,’ ‘sex at birth,’ and ‘gender’ interchangeably throughout the article.

The CDC reports that Blacks demonstrate a twofold increased risk of MM compared to Whites.^5,6^ This association could reflect environmental risk factors that are more prevalent in the Black population, as well as genetic risk factors. The CDC also reports that men are 50% more likely A few years ago, the National Institutes of Health (NIH) introduced the “All of Us” longitudinal cohort study to amass a diverse biomedical dataset from one million or more participants across the U.S.^7^ As of February 2024, there are 413,457 participants in this study, among whom 245,394 have had their genome sequenced. Using a multivariable analysis, we assessed the different risk factors for MM diagnosis across genetic ancestry, demographics, health insurance status, body mass index, social determinants of health, and pre-existing common comorbidities. Recent advances in high-throughput whole genome sequencing have confirmed the fundamental importance of epistatic effects, or genetic interactions where the effect of a genetic factor on a phenotype is conditional to the presence of others. In this study, we hypothesize that race and certain complex diseases or traits, such as hypertension and diabetes, may have epistatic effects or interactions due to their influence on genetic background and susceptibility. These interactions should be considered to understand better and identify risk factors for multiple myeloma (MM).

## METHODS

### Overview

This study is a retrospective analysis of all participants in the All of Us Controlled Tier Dataset v7, released in April 2023. The All of Us data provides information obtained through electronic health records (including conditions, drug exposures, labs & measurements, and procedures), surveys (including demographics, e.g., age, sex, and race, lifestyle, and overall health), physical measurements (e.g., body mass index), genetic data, socioeconomic status (SES) summary statistics sourced from the U.S. Census American Community Survey via a three-digit zip code linkage. Each table is linked to each other through unique participant identification numbers. This retrospective study did not require IRB approval.

### Participants

There were 413,457 participants, of whom 1,430 developed a diagnosis of plasma cell myeloma (1,414 participants, SNOMED 109989006) or plasmacytoma (16 participants, SNOMED 415112005).

### Outcomes

The main outcome is the diagnosis of multiple myeloma, as reported in the “condition” table of the database.

### Covariates

In our analysis, the model incorporated several covariates. To identify the covariates in the All of Us databases, we reviewed and selected each independent “standard concept name” (See our appendix). These included ancestry genetic data derived from participants’ exomes, demographic variables: age, race, and sex at birth; the Body Mass Index (BMI); common comorbidities: congestive heart failure (CHF), chronic obstructive pulmonary disease (COPD), hypertension, and diabetes; and social determinants of health, captured by the zip code deprivation index and self-reported health insurance status. For participants diagnosed with MM, since our study focuses on identifying risk factors, we only considered comorbidities diagnosed before their MM diagnosis. For gender, we used participants’ chromosomes data (XY or XX). When it was not available, we used self-reported sex at birth or gender. For BMI, we used the most recent measurement recorded within the 12 months following the MM diagnosis, assuming that a patient’s weight is not significantly impacted by the initial MM diagnosis up to 12 months. If the BMI was unavailable, we computed it using the height and weight when available (the weight must have been measured no later than 12 months after the MM diagnosis). Finally, we classified the BMI into 4 categories (underweight: up to 18.5, normal: 18.5 to 25, overweight: 25 to 30, obese: above 30). In the case of hypertension, identification was based not only on a formal diagnosis from the “condition” database but also on the use of antihypertensive drugs or the average of the last three blood pressure measurements (if both diastolic and systolic pressures indicate hypertension by being respectively above 80 mmHg and 130 mmHg). Similarly, diabetes was identified through a formal diagnosis, using diabetes drugs, or by analyzing glucose levels (A level above 200 mg/dL would indicate diabetes) and HbA1c measurements (An HbA1c measurement above 6.5% would indicate diabetes). On the other hand, COPD and CHF were strictly identified through formal diagnoses. To assess patients’ social determinants of health, we used their zipcode’s community deprivation index and self-reported health insurance.^8^ The deprivation index is a composite index created in 2019 to quantify a community’s deprivation. It is derived from five US Census metrics (education, income, poverty, housing, and healthcare access). We categorized the zip code deprivation index into two categories, below and above the 2018 US deprivation index’s median. Health insurance status was determined using the “survey” database when available. If this information was not available, we employed a probabilistic imputation method. This method estimated an individual’s health insurance status based on the percentage of people with health insurance in their zip code, allowing us to fill in missing data with a statistically informed guess. Finally, to avoid reporting biases, we required that participants included in the analysis had been followed for at least four months.

### Validation and quality control of the All of Us database and our methods

We performed two validation tests of the database and our methods. First, we ensured that the relative risk factors of age and race reported by the CDC at the national level are consistent with the same relative risks in the All of Us database (Table 4). Second, we conducted the same multivariate analysis using a negative control: the diagnosis of an open fracture. We chose open fractures as our negative control because, unlike closed fractures that may result from comorbidities such as osteoporosis or cancer metastasis, open fractures are typically caused by accidents and random events. Therefore, they are less likely to be influenced by pre-existing comorbidities. Finally, a Variance Inflation Factor (VIF) analysis, commonly used to detect colinearity between variables, shows that all the variables included in the model have VIF values from 1.0 to 2.0, well below the common threshold of 5, which suggests no concerning level of multicollinearity among these predictors.

### Statistical analysis

Demographic and clinical characteristics were described and then stratified by genetic ancestry (Table 2) and MM diagnosis (Table 3). Bivariate analyses between race (Table 2) or MM diagnosis (Table 3) and clinical-demographic characteristics were performed using the chi-square (χ^2^) statistical test for categorical variables and the (non-parametric) Kruskal–Wallis test for continuous variables after checking that these parametric test requirements were met. These analyses were conducted after the data met the assumptions necessary for the chi-square test, such as expected frequency distribution and sample size adequacy. Multivariable logistic regression models were conducted to identify factors independently associated with being diagnosed with multiple myeloma, first adjusted for age, self-reported race/ethnicity, and sex to ensure that the All of Us database reflects CDC-reported national trends (Table 4), then adjusted for all covariates but we replaced self-reported race/ethnicity with predominant genetic ancestry (Table 5), and finally adjusted for all covariates with interactions driven by the predominant genetic ancestry (Table 6).

**Table 1.** Distribution of participants with and without MM by self-reported race/ethnicity. Standard errors for each percentage are included. The p-value was calculated using a chi-square test.

|  | White | Black | Hispanic | Total | p-value |
| --- | --- | --- | --- | --- | --- |
| **Not diagnosed with MM** | 102,394 (99.71% ± 0.02%) | 34,193 (99.61% ± 0.03%) | 29,724 (99.80% ± 0.03%) | 166,311 (99.71% ± 0.01%) | < 1e-3 |
| **Diagnosed with MM** | 298 (0.29% ± 0.02%) | 135 (0.39% ± 0.03%) | 59 (0.20% ± 0.03%) | 492 (0.29% ± 0.01%) | < 1e-3 |
| **Total** | 102,692 | 34,328 | 29,783 | 166,803 |  |

**Table 2.** Study participant characteristics by predominant genetic ancestry with their respective percentages and standard errors on their proportions. The p-value was calculated using a Kruskal–Wallis test for the age and a chi-square test for all other variables.

|  | European | African | Ad Mixed American | Total | P-value |
| --- | --- | --- | --- | --- | --- |
| **MM diagnosis** |  |  |  |  |  |
| MM | 312<br>(0.3% ± 0.0%) | 139<br>(0.4% ± 0.0%) | 41<br>(0.2% ± 0.0%) | 492<br>(0.29% ± 0.0%) | <1e-3 |
| **Demographics** |  |  |  |  |  |
| Age (mean ± std) | 57.9 (± 16.9) | 52.0 (± 14.8) | 47.5 (± 15.7) | 55.2 (± 16.8) | <1e-3 |
| Female | 67,278<br>(62.4% ± 0.1%) | 22,751<br>(62.2% ± 0.3%) | 15,356<br>(68.7% ± 0.3%) | 105,385<br>(63.18% ± 0.1%) | <1e-3 |
| **BMI Group** |  |  |  |  |  |
| Underweight | 1,280<br>(1.2% ± 0.0%) | 544<br>(1.5% ± 0.1%) | 188<br>(0.8% ± 0.1%) | 2,012<br>(1.21% ± 0.0%) | <1e-3 |
| Normal | 30,834<br>(28.6% ± 0.1%) | 7,612<br>(20.8% ± 0.2%) | 4,144<br>(18.5% ± 0.3%) | 42,590<br>(25.53% ± 0.1%) | <1e-3 |
| Overweight | 34,675<br>(32.1% ± 0.1%) | 9,657<br>(26.4% ± 0.2%) | 6,895<br>(30.9% ± 0.3%) | 51,227<br>(30.71% ± 0.1%) | <1e-3 |
| Obese | 41,101<br>(38.1% ± 0.1%) | 18,750<br>(51.3% ± 0.3%) | 11,123<br>(49.8% ± 0.3%) | 70,974<br>(42.55% ± 0.1%) | <1e-3 |
| **Comorbidities** |  |  |  |  |  |
| Hypertension | 70,188<br>(65.1% ± 0.1%) | 26,350<br>(72.1% ± 0.2%) | 12,532<br>(56.1% ± 0.3%) | 109,070<br>(65.39% ± 0.1%) | <1e-3 |
| Diabetes | 49,730<br>(46.1% ± 0.2%) | 19,286<br>(52.7% ± 0.3%) | 10,571<br>(47.3% ± 0.3%) | 79,587<br>(47.71% ± 0.1%) | <1e-3 |
| CHF | 5,685<br>(5.3% ± 0.1%) | 2,896<br>(7.9% ± 0.1%) | 818<br>(3.7% ± 0.1%) | 9,399<br>(5.63% ± 0.1%) | <1e-3 |
| COPD | 9,151<br>(8.5% ± 0.1%) | 3,908<br>(10.7% ± 0.2%) | 900<br>(4.0% ± 0.1%) | 13,959<br>(8.37% ± 0.1%) | <1e-3 |
| **Social Determinants of Health** |  |  |  |  |  |
| No health insurance | 11,279<br>(10.5% ± 0.1%) | 5,399<br>(14.8% ± 0.2%) | 4,144<br>(18.5% ± 0.3%) | 20,822<br>(12.48% ± 0.1%) | <1e-3 |
| Deprivation index above national median | 23,762<br>(22.0% ± 0.1%) | 18,348<br>(50.2% ± 0.3%) | 10,813<br>(48.4% ± 0.3%) | 52,923<br>(31.73% ± 0.1%) | <1e-3 |

**Table 3.**
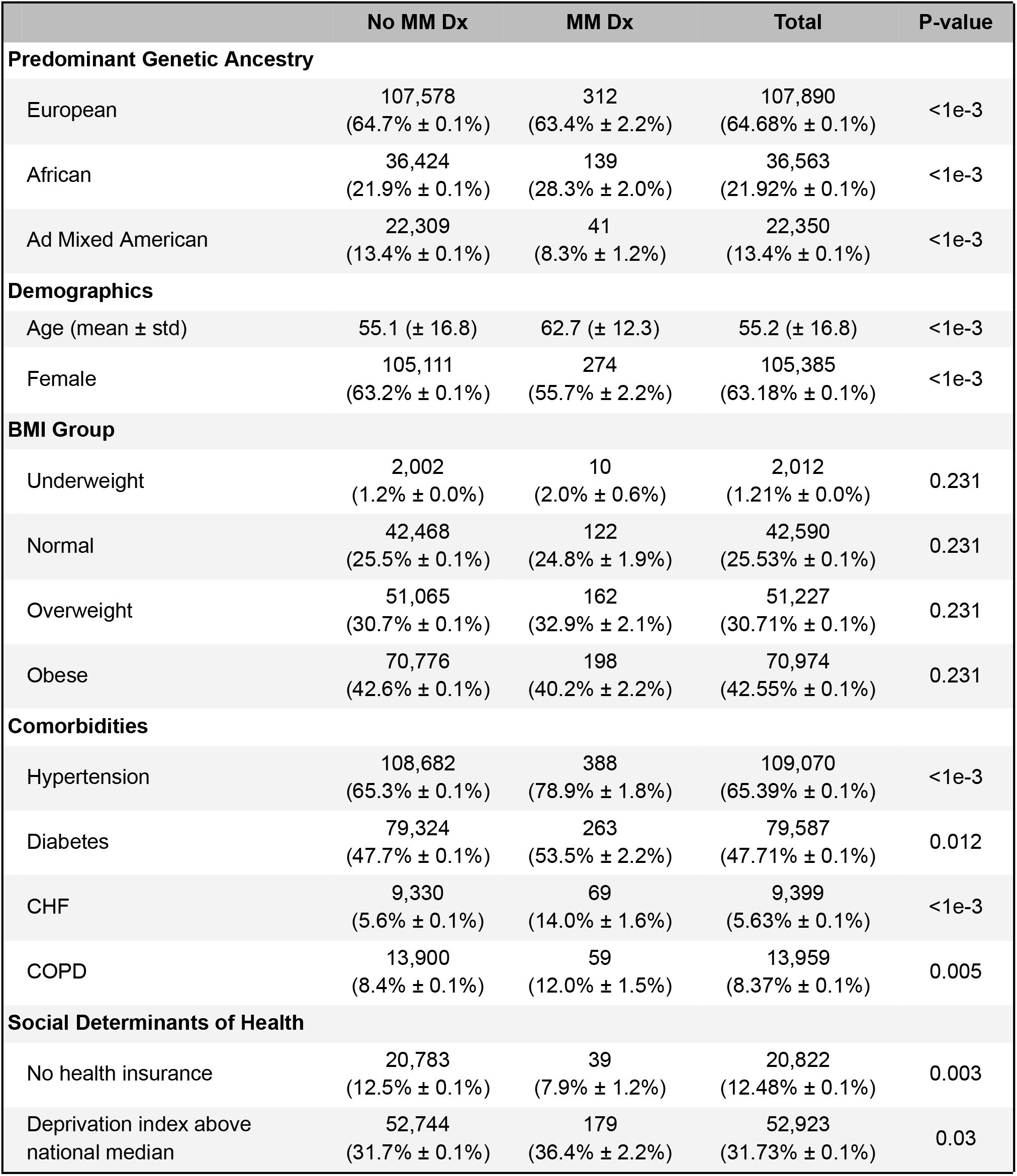
Study participant characteristics by MM diagnosis with their respective percentages and standard errors on their proportions. The p-value was calculated using a Kruskal–Wallis test for the age and a chi-square test for all other variables.

## RESULTS

### Characteristics of the study population

In this dataset, there are 413,457 individuals, of whom 1,430 (0.35%) were diagnosed with MM. We only retained 166,803 participants, corresponding to roughly 40% of the original dataset; 492 were diagnosed with MM. We excluded participants who lacked genetic data, gender, zip code, BMI, or for whom All of Us had less than four months of data. To ensure that the resulting dataset was representative of the US population, we conducted a multivariable analysis on the dataset, controlling for age, self-reported race/ethnicity, and gender, following CDC standards. We verified that the odds ratios aligned closely with those published by the CDC (See Table 4). Of the remaining 168,290 participants, the mean age was 55.2 years old (Standard Deviation: 16.8 years), and 63.18% were women (Standard Error: 0.1%). Table 1 shows the proportion of MM participants by self-reported race/ethnicity. “Black” refers to non-Hispanic Black, and “White” refers to non-Hispanic “White.” “Hispanic” refers to people reporting a “Hispanic” ethnicity without reporting race.

**Table 4.**
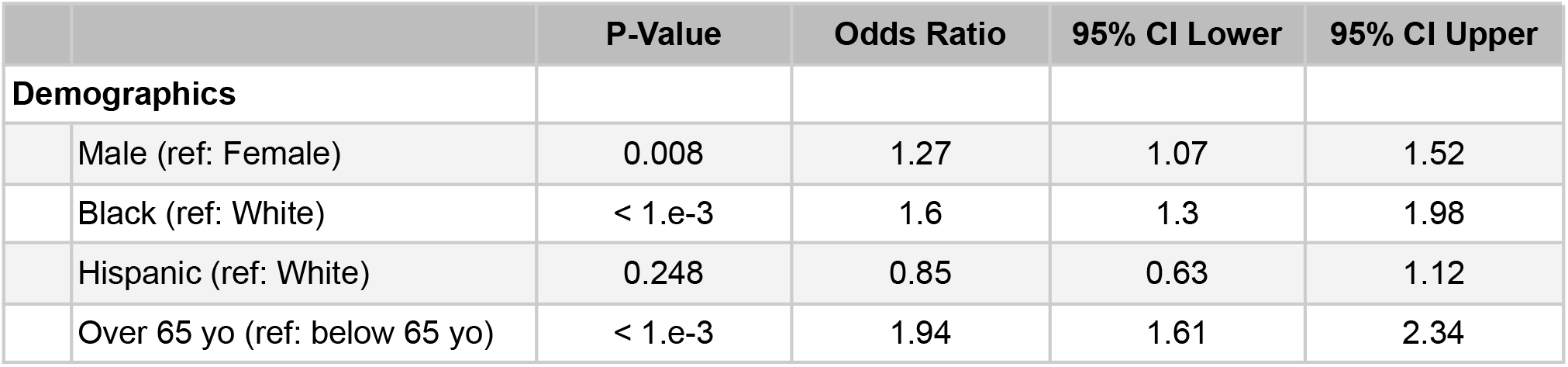
Multivariate analysis for MM risks in our dataset with self-reported race/ethnicity, gender, and age as covariates. This analysis replicates the CDC statistics and shows that our dataset is consistent with the trends observed by the CDC.

**Table 5.** Fully adjusted model for MM risk without taking into account interactions.

|  | P-Value | Odds Ratio | 95% CI Lower | 95% CI Upper |
| --- | --- | --- | --- | --- |
| **Predominant Genetic Ancestry** |  |  |  |  |
| African (ref: European) | 0.005 | 1.36 | 1.1 | 1.69 |
| Ad Mixed American (ref: European) | 0.127 | 0.77 | 0.55 | 1.08 |
| **Demographics** |  |  |  |  |
| Male (ref: Female) | 0.043 | 1.21 | 1.01 | 1.44 |
| Over 65 yo (ref: below 65 yo) | < 1.e-3 | 1.63 | 1.35 | 1.98 |
| **BMI Group (ref: BMI in normal range)** |  |  |  |  |
| Underweight | 0.097 | 1.73 | 0.9 | 3.31 |
| Overweight | 0.923 | 0.99 | 0.78 | 1.25 |
| Obese | 0.186 | 0.85 | 0.68 | 1.08 |
| **Comorbidities** |  |  |  |  |
| Hypertension (ref: No Hypertension) | < 1.e-3 | 1.63 | 1.28 | 2.07 |
| Diabetes (ref: No Diabetes) | 0.372 | 0.91 | 0.75 | 1.11 |
| CHF (ref: No CHF) | < 1.e-3 | 2.08 | 1.58 | 2.75 |
| COPD (ref: No COPD) | 0.786 | 0.96 | 0.72 | 1.28 |
| **Social Determinants of Health** |  |  |  |  |
| Not insured (ref: insured) | 0.015 | 0.66 | 0.48 | 0.92 |
| Deprivation index above national median (ref: below) | 0.019 | 1.26 | 1.04 | 1.53 |

### Characteristics of participants included in analyses

Table 2 shows how demographics, BMI, comorbidities, and social determinants of health differ among the three main racial/ethnic groups. Table 3 shows how these variables differ among participants diagnosed or not with MM.

### Model adjusted for age, sex, and race

In Table 4, as a quality control of our methods, we report a first-approach multivariate analysis to assess the effect of the main demographic factors (age, gender, and race/ethnicity) on MM risk. We note that these disparities are consistent with those reported by the CDC,^9^, suggesting that the dataset, after removing the missing variables, is representative of the US population and does not suffer from major selection bias.

### Predominant Genetic Ancestry

In Figure 1, we present a principal component analysis (PCA) using participants’ exome data and a reference model trained on the 1,000 Genome Project genotype data. This approach is designed to cluster genomes based on their predominant genetic ancestry in a multi-dimensional space where the principal components are built to account for the greatest variance and effectively separate populations according to their genetic similarities and differences. We used the five “super populations” to color the data points: African, Ad Mixed American, East Asian, European, and South Asian.^10^ We present two two-dimensional representations of the PCA, with and without hypertension, and an overlay of the MM diagnosis. As demonstrated by our fully-adjusted multivariate model with interactions (See Table 6), the MM cluster overlaying the African population is much denser in participants with hypertension compared to those without hypertension. Unlike our multivariate analysis, which included only participants with complete datasets, this PCA includes all participants with available genetic data, even if other types of data (e.g., BMI) are missing.

**Table 6.**
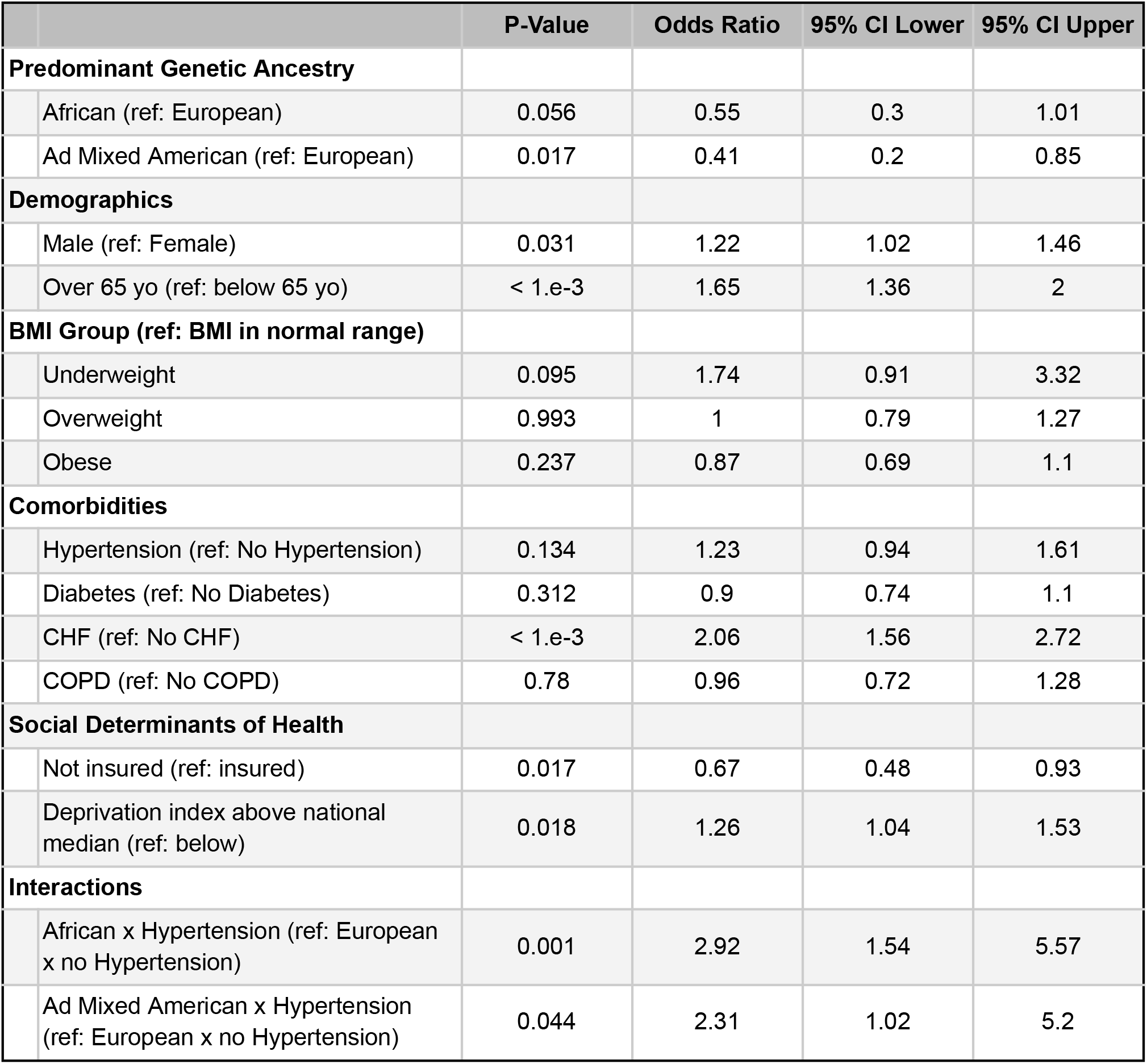
Fully-adjusted model for MM risk with interactions (predominant genetic ancestry x hypertension)

**Figure 1.**
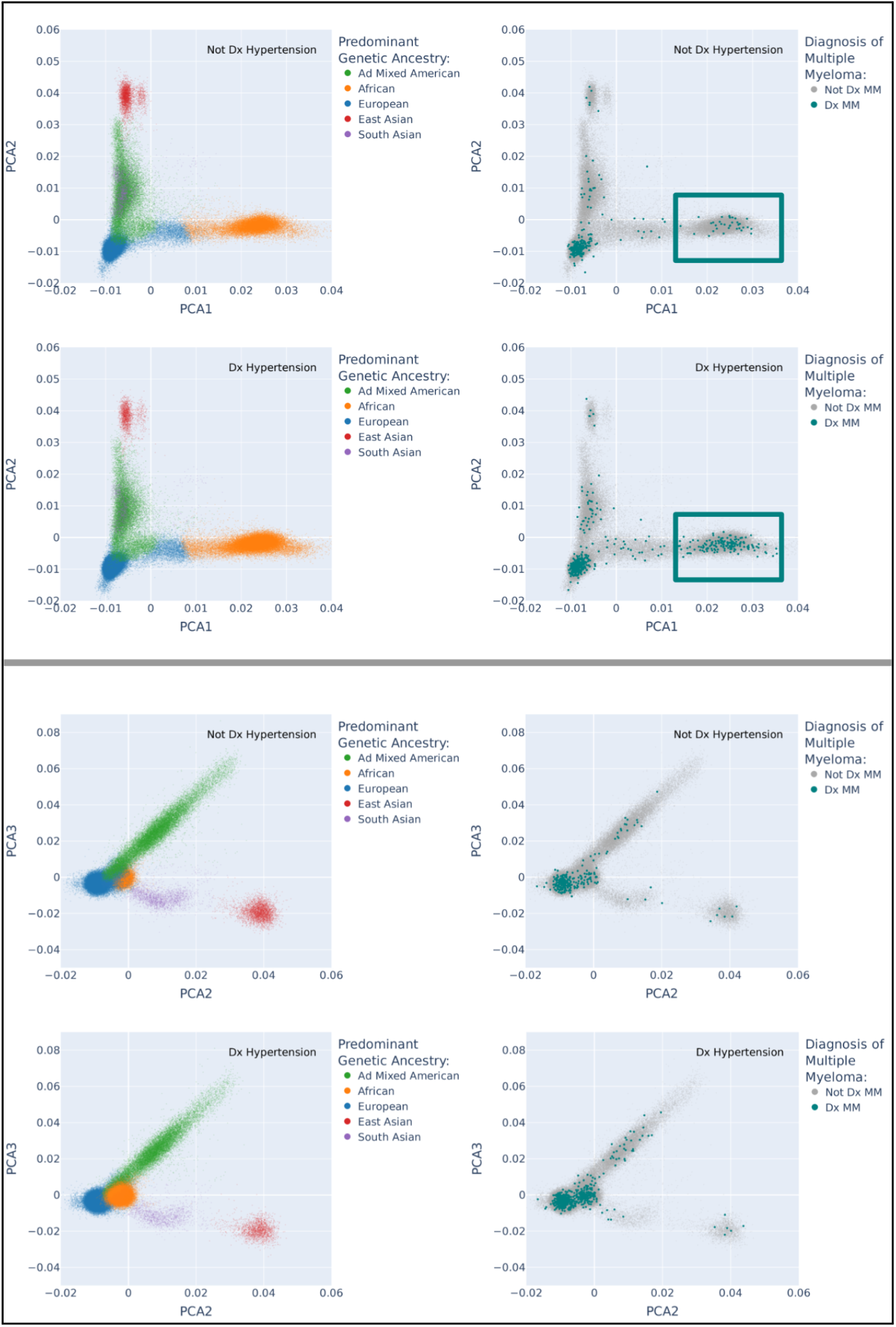
Principal Component Analysis (PCA) displaying predominant genetic ancestry, hypertension diagnosis, and Multiple Myeloma (MM) diagnosis. Number of patients without MM: 244,522. Number of patients with MM: 866. The two green squares represent the difference in MM density in participants who have a predominantly African genetic ethnicity with and without hypertension. The Genetic Ethnicity was obtained using a reference model trained on the 1,000 Genome Project genotype data. Unlike our multivariate analysis, which included only participants with complete datasets, this PCA includes all participants with available genetic data, even if other types of data (e.g., BMI) are missing.

### Fully-adjusted model without interaction

We then conducted a fully adjusted multivariate analysis without interactions, which is reported in Table 5. Even after adjusting for demographics, comorbidities, and social determinants of health, the association between MM and predominant genetic ancestry is significant. Surprisingly, hypertension and CHF are also significantly associated with MM risk (Table 5), prompting us to look further into these comorbidities and their potential interaction with the predominant genetic ancestry.

### Fully-adjusted model with race-driven interactions

We constructed a multivariable model that revealed significant interactions between the predominant genetic ancestry and hypertension. We also included an interaction between predominant genetic ancestry and CHF, but it was not significant (not shown). We conducted a stratified analysis to better understand the relationship between predominant genetic ancestry and hypertension and their association with MM. In the tables stratified by gender, we included the interaction between predominant genetic ancestry and hypertension. The Appendix shows these results: Tables S1, S2, and S3 represent the risk factor model for the European, African, and AD Mixed American populations, respectively. Tables S4 and S5 represent the risk factor model for the Male and Female populations, respectively. We then estimated and compared the odds ratios of each combination. Using Table S4, we found that males whose predominant genetic ancestry is African and without hypertension were at a significantly lower risk of MM compared to Europeans without hypertension (OR: 0.32, 95% CI: 0.11 to 0.9, P = 0.03), while males whose predominant genetic ancestry is African and with hypertension have a significantly increased risk compared to Europeans and with hypertension (OR: 3.35, 95% CI: 1.13 to 9.99, P = 0.03). Using Table S5, we found that females whose predominant genetic ancestry is African and without hypertension were not at significant risk of MM compared to European females without hypertension (OR: 0.8, 95% CI: 0.37 to 1.72, P = 0.568), while females whose predominant genetic ancestry is African with hypertension were at significant risk of MM compared to European females with hypertension (OR: 2.7, 95% CI: 1.2 to 6.04, P = 0.016). This goes to show that male participants whose predominant genetic ancestry is African have genetic protection against MM when they do not have hypertension. Table S1 also shows that participants (male and female) whose predominant genetic ancestry is European and having been diagnosed with hypertension were not at higher risk for MM (OR: 1.25, 95% CI: 0.94 to 1.65, P = 0.12). Table S2 and S3 also shows that participants whose predominant genetic ancestry is African or Ad Mixed American and with hypertension are at higher risk than those without hypertension (OR: 3.44, 95% CI: 1.84 to 6.42, P = < 1.e-3 and OR: 2.82, 95% CI: 1.2 to 6.63, P = 0.018 respectively). Our stratified analysis confirms that hypertension is an important risk factor for participants whose predominant genetic ancestry is African or Ad Mixed American, but not for Europeans.

## DISCUSSION

Our investigation delved into the racial/ethnic inequalities observed in the likelihood of receiving an MM diagnosis by the CDC. We aimed to discern whether such disparities can be elucidated through genetics, demographics, BMI, social determinants of health, and common pre-existing health conditions. Building on the well-documented racial disparity between Whites and Blacks from the CDC data, our analysis suggests that the elevated risk of Black individuals for MM stems from an epistatic interaction between genetic African ethnicity and having hypertension (OR: 2.92, 95% CI: 1.54 to 5.57, P = 0.001). In our final model (Table 6), hypertension is not a significant risk factor for the European population (OR: 1.23, 95% CI: 0.94 to 1.61, P = 0.134). Other studies have shown that MM patients are more likely to be diagnosed with hypertension, confirming the results of our fully adjusted model without the interaction between hypertension and predominant genetic ancestry (Table 5).^11^

When running an univariate model between MM and BMI (See appendix), we did not find a significant association between being overweight and an increased risk of MM. Also, our final model (Table 6) shows no significant association between BMI and MM risk. However, several previous studies showed a positive association between obesity and the MM death rate.^12–14^ This discrepancy could stem from several reasons: the sample population from the “All of Us” study differs from those in previous studies in terms of how participants were selected, or the mortality rate, a focus of previous studies, does not strongly correlate with disease risk, which was the main focus of our study.

Our analysis also recognized CHF as a significant risk factor (OR: 2.06, 95% CI: 1.56 to 2.72, P < 1.e-3). Cardiovascular issues have been associated with MM patients throughout their treatment^15^ but are not a known risk for MM diagnosis. Although hypertension and CHF might share some environmental and genetic risk factors, our adjusted analysis suggests an independent effect of CV. However, similar to hypertension, one explanation for this association could be the presence of genetic variants associated with both CHF and MM. Further work is required to test this association and the role of genetic susceptibility.

An another dimension added to our study was the substantial link we identified between social determinants of health, gauged via health insurance status and the zip code deprivation index, and the likelihood of an MM diagnosis. All of Us participants living in deprived areas are 26% more likely to be diagnosed with MM (OR: 1.26, 95% CI: 1.04 to 1.53, P = 0.018). Earlier research on the role of social deprivation in cancer susceptibility and progression has yielded mixed results. For instance, Rosenzweig et al. associated area deprivation with heightened anxiety levels in advanced cancer patients,^16^ whereas Luningham et al. showed that breast cancer in the state of Georgia was associated with race and not the environment.^17^ Meanwhile, Cheng et al. showed that the environment is significantly associated with survival among patients with nonmetastatic common cancers.^18^ Furthermore, a comprehensive study with a cohort exceeding one million participants indicated a significant association between area deprivation and elevated lung cancer prevalence and mortality, even after adjusting for rurality.^19^ More work, however, is needed to delineate what environmental factors increase someone’s risk for MM (exposure to pollutants, dietary habits, etc.). Lastly, in line with existing literature, our analysis found that uninsured participants had a 33% reduced probability of receiving an MM diagnosis (OR: 0.67, 95% CI: 0.48 to 0.93, P = 0.017). Given that uninsured individuals tend to access medical care less frequently compared to those with insurance, a greater percentage of uninsured individuals might be living with undiagnosed MM, which can be an indolent disease in the initial stages. This trend aligns with the observation that uninsured patients are often diagnosed with late-stage cancer, which consequently results in lower survival rates when compared to insured individuals.^20^

### Strengths and limitations

The main strength of our study is the large sample size made available by the ‘All of Us’ database. Unlike prior studies, our model adjusts for all genomic and non-genomic factors. The main limitation of this study is the possible bias in All of Us’ enrollment, which excludes people who did not want to participate or were unaware they could participate. A second limitation is that there may be some residual confounding that was not taken into account by our model.

## CONCLUSIONS

Our research provides novel insights into the racial disparities related to MM diagnosis. We found that the discrepancy in MM incidence between White and Black individuals, as documented by the CDC, results from an interplay between having African as a predominant genetic ancestry and hypertension. Consequently, hypertension can serve as an indicator to identify individuals with African predominant genetic ancestry who are at higher risk for MM. This novel finding paves the way for future studies to develop a precise genetic panel for identifying individuals at high risk for MM.

## Data Availability

All data produced in the present study are available upon reasonable request to the authors

https://workbench.researchallofus.org/

## ACKNOWLEDGEMENTS

The All of Us Research Program is supported by the National Institutes of Health, Office of the Director: Regional Medical Centers: 1 OT2 OD026549; 1 OT2 OD026554; 1 OT2 OD026557; 1 OT2 OD026556; 1 OT2 OD026550; 1 OT2 OD 026552; 1 OT2 OD026553; 1 OT2 OD026548; 1 OT2 OD026551; 1 OT2 OD026555; IAA #: AOD 16037; Federally Qualified Health Centers: HHSN 263201600085U; Data and Research Center: 5 U2C OD023196; Biobank: 1 U24 OD023121; The Participant Center: U24 OD023176; Participant Technology Systems Center: 1 U24 OD023163; Communications and Engagement: 3 OT2 OD023205; 3 OT2 OD023206; and Community Partners: 1 OT2 OD025277; 3 OT2 OD025315; 1 OT2 OD025337; 1 OT2 OD025276. In addition, the All of Us Research Program would not be possible without the partnership of its participants. We thank the Hackensack Meridian Center for Discovery and Innovation and the BD Foundation for their generous funding.

## REFERENCES

1. Cowan, A. J. et al. Diagnosis and Management of Multiple Myeloma: A Review. JAMA 327, 464–477 (2022).

2. Peres, L. C., Hansen, D. K., Maura, F. & Kazandjian, D. The knowns and unknowns of disparities, biology, and clinical outcomes in Hispanic and Latinx multiple myeloma patients in the U.S. Semin. Oncol. 49, 3–10 (2022).

3. Mateos, M.-V. et al. Global disparities in patients with multiple myeloma: a rapid evidence assessment. Blood Cancer J. 13, 109 (2023).

4. Howlader, N., Noone, A. M. & Krapcho, M. SEER cancer statistics review, 1975–2012. National Cancer (2014).

5. SEER*Explorer. https://seer.cancer.gov/statistics-network/explorer/application.html?site=89&data_type=1&graph_type=2&compareBy=race&chk_race_6=6&chk_race_5=5&chk_race_4=4&chk_race_9=9&chk_race_8=8&rate_type=2&sex=1&age_range=1&stage=101&advopt_precision=1&advopt_show_ci=on&hdn_view=0&advopt_show_apc=on&advopt_display=1.

6. Marinac, C. R., Ghobrial, I. M., Birmann, B. M., Soiffer, J. & Rebbeck, T. R. Dissecting racial disparities in multiple myeloma. Blood Cancer J. 10, 19 (2020).

7. Ramirez, A. H., Gebo, K. A. & Harris, P. A. Progress With the All of Us Research Program: Opening Access for Researchers. JAMA 325, 2441–2442 (2021).

8. Brokamp, C. et al. Material community deprivation and hospital utilization during the first year of life: an urban population-based cohort study. Ann. Epidemiol. 30, 37–43 (2019).

9. SEER*Explorer. https://seer.cancer.gov/statistics-network/explorer/application.html?site=89&data_type=1&graph_type=2&compareBy=sex&chk_sex_3=3&chk_sex_2=2&rate_type=2&race=1&age_range=1&stage=101&advopt_precision=1&advopt_show_ci=on&hdn_view=0&advopt_show_apc=on&advopt_display=1.

10. Help - Frequently Asked Questions - Homo_sapiens - Ensembl genome browser 112. https://useast.ensembl.org/Help/Faq?id=532.

11. Chari, A. et al. Incidence and risk of hypertension in patients newly treated for multiple myeloma: a retrospective cohort study. BMC Cancer 16, 912 (2016).

12. Marques-Mourlet, C., Di Iorio, R., Fairfield, H. & Reagan, M. R. Obesity and myeloma: Clinical and mechanistic contributions to disease progression. Front. Endocrinol. 14, 1118691 (2023).

13. Lichtman, M. A. Obesity and the risk for a hematological malignancy: leukemia, lymphoma, or myeloma. Oncologist 15, 1083–1101 (2010).

14. Calle, E. E., Rodriguez, C., Walker-Thurmond, K. & Thun, M. J. Overweight, obesity, and mortality from cancer in a prospectively studied cohort of U.S. adults. N. Engl. J. Med. 348, 1625–1638 (2003).

15. El-Cheikh, J., Moukalled, N., Malard, F., Bazarbachi, A. & Mohty, M. Cardiac toxicities in multiple myeloma: an updated and a deeper look into the effect of different medications and novel therapies. Blood Cancer J. 13, 83 (2023).

16. Rosenzweig, M. Q. et al. The Association Between Area Deprivation Index and Patient-Reported Outcomes in Patients with Advanced Cancer. Health Equity 5, 8–16 (2021).

17. Luningham, J. M. et al. Association of Race and Area Deprivation With Breast Cancer Survival Among Black and White Women in the State of Georgia. JAMA Netw Open 5, e2238183 (2022).

18. Cheng, E. et al. Neighborhood and Individual Socioeconomic Disadvantage and Survival Among Patients With Nonmetastatic Common Cancers. JAMA Netw Open 4, e2139593 (2021).

19. Fairfield, K. M. et al. Area Deprivation Index and Rurality in Relation to Lung Cancer Prevalence and Mortality in a Rural State. JNCI Cancer Spectr 4, kaa011 (2020).

20. Dwyer, L. L. et al. Disparities in Lung Cancer: A Targeted Literature Review Examining Lung Cancer Screening, Diagnosis, Treatment, and Survival Outcomes in the United States. J Racial Ethn Health Disparities (2023) doi:10.1007/s40615-023-01625-2.

