## Supplementary Information for "Genetic Ethnicity and Hypertension Epistatic Interaction Underlying Racial Disparities in US Multiple Myeloma Susceptibility"

Appendix

| PREDOMINANT GENETIC ANCESTRY: EUROPEAN | | P-Value | Odds Ratio | 95% CI Lower | 95% CI Upper |
|---|---|---|---|---|---|
| **Demographics** | | | | | |
| | Male (ref: Female) | < 1.e-3 | 1.57 | 1.25 | 1.97 |
| | Over 65 yo (ref: below 65 yo) | < 1.e-3 | 1.73 | 1.36 | 2.19 |
| **BMI Group (ref: BMI in normal range)** | | | | | |
| | Underweight | 0.458 | 1.41 | 0.57 | 3.48 |
| | Overweight | 0.743 | 0.95 | 0.72 | 1.26 |
| | Obese | 0.058 | 0.75 | 0.56 | 1.01 |
| **Comorbidities** | | | | | |
| | Hypertension (ref: No Hypertension) | 0.12 | 1.25 | 0.94 | 1.65 |
| | Diabetes (ref: No Diabetes) | 0.095 | 0.81 | 0.63 | 1.04 |
| | CHF (ref: No CHF) | 0.001 | 1.85 | 1.27 | 2.69 |
| | COPD (ref: No COPD) | 0.352 | 1.18 | 0.83 | 1.69 |
| **Social Determinants of Health** | | | | | |
| | Not insured (ref: insured) | 0.103 | 0.7 | 0.45 | 1.08 |
| | Deprivation index above national median (ref: below) | 0.003 | 1.45 | 1.14 | 1.86 |

**Table S1. Risk factor analysis for the European population**

| PREDOMINANT GENETIC ANCESTRY: AFRICAN | | P-Value | Odds Ratio | 95% CI Lower | 95% CI Upper |
|---|---|---|---|---|---|
| **Demographics** | | | | | |
| | Male (ref: Female) | 0.08 | 0.72 | 0.49 | 1.04 |
| | Over 65 yo (ref: below 65 yo) | 0.003 | 1.75 | 1.21 | 2.51 |
| **BMI Group (ref: BMI in normal range)** | | | | | |
| | Underweight | 0.339 | 1.81 | 0.54 | 6.08 |
| | Overweight | 0.7 | 1.11 | 0.65 | 1.89 |
| | Obese | 0.823 | 1.06 | 0.65 | 1.73 |
| **Comorbidities** | | | | | |
| | Hypertension (ref: No Hypertension) | < 1.e-3 | 3.44 | 1.84 | 6.42 |
| | Diabetes (ref: No Diabetes) | 0.552 | 0.89 | 0.61 | 1.3 |
| | CHF (ref: No CHF) | < 1.e-3 | 2.59 | 1.66 | 4.03 |
| | COPD (ref: No COPD) | 0.257 | 0.74 | 0.44 | 1.24 |
| **Social Determinants of Health** | | | | | |
| | Not insured (ref: insured) | 0.241 | 0.71 | 0.4 | 1.26 |
| | Deprivation index above national median (ref: below) | 0.573 | 1.1 | 0.79 | 1.54 |

**Table S2. Risk factor analysis for the African population**

| PREDOMINANT GENETIC ANCESTRY: AD MIXED AMERICAN | | P-Value | Odds Ratio | 95% CI Lower | 95% CI Upper |
|---|---|---|---|---|---|
| **Demographics** | | | | | |
| | Male (ref: Female) | 0.49 | 1.25 | 0.66 | 2.37 |
| | Over 65 yo (ref: below 65 yo) | 0.371 | 0.67 | 0.27 | 1.62 |
| **BMI Group (ref: BMI in normal range)** | | | | | |
| | Underweight | 0.015 | 6.98 | 1.45 | 33.63 |
| | Overweight | 0.452 | 0.7 | 0.28 | 1.76 |
| | Obese | 0.387 | 0.69 | 0.3 | 1.59 |
| **Comorbidities** | | | | | |
| | Hypertension (ref: No Hypertension) | 0.018 | 2.82 | 1.2 | 6.63 |
| | Diabetes (ref: No Diabetes) | 0.111 | 1.8 | 0.87 | 3.72 |
| | CHF (ref: No CHF) | 0.578 | 1.41 | 0.42 | 4.76 |
| | COPD (ref: No COPD) | 0.301 | 0.34 | 0.05 | 2.59 |
| **Social Determinants of Health** | | | | | |
| | Not insured (ref: insured) | 0.19 | 0.5 | 0.18 | 1.41 |
| | Deprivation index above national median (ref: below) | 0.846 | 0.94 | 0.51 | 1.74 |

**Table S3. Risk factor analysis for the AD Mixed American population**

| GENDER: MALE | | P-Value | Odds Ratio | 95% CI Lower | 95% CI Upper |
|---|---|---|---|---|---|
| **Predominant Genetic Ancestry** | | | | | |
| | African (ref: European) | 0.03 | 0.32 | 0.11 | 0.9 |
| | Ad Mixed American (ref: European) | 0.115 | 0.39 | 0.12 | 1.26 |
| **BMI Group (ref: BMI in normal range)** | | | | | |
| | Underweight | 0.089 | 2.44 | 0.87 | 6.83 |
| | Overweight | 0.955 | 0.99 | 0.69 | 1.41 |
| | Obese | 0.945 | 0.99 | 0.69 | 1.42 |
| **Comorbidities** | | | | | |
| | Hypertension (ref: No Hypertension) | 0.623 | 0.91 | 0.61 | 1.34 |
| | Diabetes (ref: No Diabetes) | 0.451 | 0.89 | 0.66 | 1.2 |
| | CHF (ref: No CHF) | 0.001 | 1.92 | 1.29 | 2.85 |
| | COPD (ref: No COPD) | 0.468 | 1.16 | 0.78 | 1.73 |
| **Social Determinants of Health** | | | | | |
| | Not insured (ref: insured) | 0.441 | 0.84 | 0.53 | 1.32 |
| | Deprivation index above national median (ref: below) | 0.192 | 1.22 | 0.91 | 1.64 |
| **Interactions** | | | | | |
| | African x Hypertension (ref: European x no Hypertension) | 0.03 | 3.35 | 1.13 | 9.99 |
| | Ad Mixed American x Hypertension (ref: European x no Hypertension) | 0.243 | 2.18 | 0.59 | 8.06 |

**Table S4. Risk factor analysis for the Male population**

| GENDER: FEMALE | | P-Value | Odds Ratio | 95% CI Lower | 95% CI Upper |
|---|---|---|---|---|---|
| **Predominant Genetic Ancestry** | | | | | |
| | African (ref: European) | 0.568 | 0.8 | 0.37 | 1.72 |
| | Ad Mixed American (ref: European) | 0.096 | 0.45 | 0.18 | 1.15 |
| **BMI Group (ref: BMI in normal range)** | | | | | |
| | Underweight | 0.368 | 1.47 | 0.64 | 3.39 |
| | Overweight | 0.763 | 0.95 | 0.69 | 1.31 |
| | Obese | 0.033 | 0.71 | 0.52 | 0.97 |
| **Comorbidities** | | | | | |
| | Hypertension (ref: No Hypertension) | 0.021 | 1.55 | 1.07 | 2.25 |
| | Diabetes (ref: No Diabetes) | 0.378 | 0.89 | 0.68 | 1.16 |
| | CHF (ref: No CHF) | < 1.e-3 | 2.24 | 1.52 | 3.32 |
| | COPD (ref: No COPD) | 0.311 | 0.8 | 0.53 | 1.23 |
| **Social Determinants of Health** | | | | | |
| | Not insured (ref: insured) | 0.017 | 0.56 | 0.35 | 0.9 |
| | Deprivation index above national median (ref: below) | 0.033 | 1.32 | 1.02 | 1.7 |
| **Interactions** | | | | | |
| | African x Hypertension (ref: European x no Hypertension) | 0.016 | 2.7 | 1.2 | 6.04 |
| | Ad Mixed American x Hypertension (ref: European x no Hypertension) | 0.111 | 2.33 | 0.82 | 6.62 |

**Table S5. Risk factor analysis for the Female population**

| | | P-Value | Odds Ratio | 95% CI Lower | 95% CI Upper |
|---|---|---|---|---|---|
| **Demographics** | | | | | |
| | Male (ref: Female) | < 1.e-3 | 1.8 | 1.62 | 2.01 |
| | Black (ref: White) | 0.155 | 0.9 | 0.78 | 1.04 |
| | Hispanic (ref: White) | < 1.e-3 | 0.69 | 0.58 | 0.81 |
| | Over 65 yo (ref: below 65 yo) | < 1.e-3 | 0.73 | 0.65 | 0.83 |
| **BMI Group (ref: BMI in normal range)** | | | | | |
| | Underweight | 0.234 | 1.27 | 0.86 | 1.9 |
| | Overweight | 0.001 | 0.79 | 0.69 | 0.91 |
| | Obese | < 1.e-3 | 0.71 | 0.62 | 0.81 |
| **Comorbidities** | | | | | |
| | Hypertension (ref: No Hypertension) | 0.135 | 1.1 | 0.97 | 1.25 |
| | Diabetes (ref: No Diabetes) | 0.223 | 1.08 | 0.96 | 1.21 |
| | CHF (ref: No CHF) | 0.998 | 1 | 0.79 | 1.26 |
| | COPD (ref: No COPD) | 0.55 | 1.06 | 0.88 | 1.28 |
| **Social Determinants of Health** | | | | | |
| | Not insured (ref: insured) | 0.981 | 1 | 0.87 | 1.15 |
| | Deprivation index above national median (ref: below) | 0.791 | 1.02 | 0.9 | 1.15 |

**Table S6. Risk factor analysis for the control disease (open fracture)**

| BMI group (ref: Normal) | | P-Value | Odds Ratio | 95% CI Lower | 95% CI Upper |
|---|---|---|---|---|---|
| | Underweight | 0.093 | 1.74 | 0.91 | 3.32 |
| | Overweight | 0.409 | 1.1 | 0.87 | 1.4 |
| | Obese | 0.818 | 0.97 | 0.78 | 1.22 |

**Table S7. Univariate model between MM and BMI**

| Concept | Standard Concept Names in All of Us |
|---|---|
| **Multiple Myeloma** | Multiple myeloma |
| | Multiple myeloma in remission |
| | Relapse multiple myeloma |
| | Smoldering myeloma |
| | Plasma cell leukemia |
| | Plasma cell leukemia in remission |
| | Plasma cell leukemia in relapse |
| | IgG myeloma |
| | Kappa light chain myeloma |
| | IgA myeloma |
| | Lambda light chain myeloma |
| | Plasmacytoma |
| | Extramedullary plasmacytoma |
| **Hypertension** | Malignant essential hypertension |
| | Labile essential hypertension |
| | Systolic essential hypertension |
| | Benign essential hypertension |
| | Essential hypertension |
| | Hypertensive disorder |
| | Hypertensive urgency |
| | Hypertensive emergency |
| | Renovascular hypertension |
| | Renal hypertension |

| | |
|---|---|
| | Benign hypertension |
| | Hypertensive crisis |
| | Malignant hypertension |
| | Intermittent hypertension |
| | Transient hypertension |
| | Labile systemic arterial hypertension |
| | Systolic hypertension |
| | Resistant hypertensive disorder |
| | Diastolic hypertension |
| | Labile diastolic hypertension |
| | Paroxysmal hypertension |
| | Parenchymal renal hypertension |
| | Hypertension with albuminuria |
| **Diabetes** | Insulin treated type 2 diabetes mellitus |
| | Diabetes mellitus type 2 without retinopathy |
| | Insulin dependent diabetes mellitus type 1A |
| | Diabetes mellitus associated with pancreatic disease |
| | Secondary endocrine diabetes mellitus |
| | Maturity-onset diabetes of the young, type 3 |
| | Insulin dependent diabetes mellitus type 1B |
| | Diabetes mellitus induced by non-steroid drugs without complication |
| | Pre-existing type 2 diabetes mellitus |
| | Diabetes mellitus type 1 without retinopathy |
| | Diabetes mellitus due to structurally abnormal insulin |
| | Steroid-induced diabetes |
| | Type 2 diabetes mellitus in obese |
| | Diabetes mellitus in the puerperium - baby delivered during previous episode of care |
| | Diabetes mellitus in the puerperium - baby delivered during current episode of care |
| | Diabetes mellitus associated with cystic fibrosis |
| | Latent autoimmune diabetes mellitus in adult |
| | Type 2 diabetes mellitus in nonobese |
| | Diabetes mellitus due to cystic fibrosis |
| | Diabetes mellitus |
| | Drug-induced diabetes mellitus |
| | Secondary diabetes mellitus |

| | |
|---|---|
| | Pre-existing type 1 diabetes mellitus |
| | Type 1 diabetes mellitus |
| | Diabetes mellitus due to pancreatic injury |
| | Type 2 diabetes mellitus with ulcer |
| | Diabetes mellitus associated with hormonal etiology |
| | Type 1 diabetes mellitus without complication |
| | Type II diabetes mellitus in remission |
| | Diabetes mellitus without complication |
| | Maturity-onset diabetes of the young |
| | Maturity onset diabetes of the young, type 1 |
| | Type 1 diabetes mellitus with ulcer |
| | Type 2 diabetes mellitus |
| | Type 2 diabetes mellitus without complication |
| **Congestive Heart Failure** | Hypertensive heart and renal disease with both (congestive) heart failure and renal failure |
| | Hypertensive heart disease with congestive heart failure |
| | Congestive heart failure stage C due to ischemic cardiomyopathy |
| | Acute on chronic heart failure co-occurrent with normal ejection fraction |
| | Congestive heart failure stage C |
| | Exacerbation of congestive heart failure |
| | Chronic right-sided congestive heart failure |
| | Biventricular congestive heart failure |
| | Symptomatic congestive heart failure |
| | Acute on chronic right-sided congestive heart failure |
| | Hypertensive heart and renal disease with (congestive) heart failure |
| | Chronic congestive heart failure |
| | Congestive heart failure due to left ventricular systolic dysfunction |
| | Congestive heart failure due to valvular disease |
| | Malignant hypertensive heart disease with congestive heart failure |
| | Congestive heart failure with right heart failure |
| | Hypertensive heart AND chronic kidney disease with congestive heart failure |
| | Acute exacerbation of chronic congestive heart failure |
| | Congestive heart failure |
| | Congestive rheumatic heart failure |
| | Acute congestive heart failure |

| | |
|---|---|
| | Benign hypertensive heart disease with congestive cardiac failure |
| **Chronic Obstructive Pulmonary Disease** | Moderate chronic obstructive pulmonary disease |
| | End stage chronic obstructive airways disease |
| | Chronic obstructive lung disease co-occurrent with acute bronchitis |
| | Severe chronic obstructive pulmonary disease |
| | Mild chronic obstructive pulmonary disease |
| | Acute infective exacerbation of chronic obstructive airways disease |
| | Asthma-chronic obstructive pulmonary disease overlap syndrome |
| | Chronic obstructive pulmonary disease with acute lower respiratory infection |
| | Acute exacerbation of chronic obstructive airways disease |
| | Chronic obstructive lung disease |
| | Emphysematous bleb of lung |
| | Chronic bullous emphysema |
| | Centriacinar emphysema |
| | Panacinar emphysema |
| | Interstitial emphysema of lung |
| | Bronchial atresia with segmental pulmonary emphysema |
| | Pulmonary emphysema |
| | Compensatory emphysema |
| | Allergic bronchopulmonary aspergillosis |
| | Aspirin exacerbated respiratory disease |
| | Chronic obliterative bronchiolitis |
| **Diabetes drug** | metformin |
| | sulfonylureas |
| | meglitinide |
| | Thiazolidinediones |
| | DPP-4 inhibitor |
| | GLP-1 receptor agonist |
| | SGLT2 inhibitors |
| | insulin |
| | pioglitazone |
| | glipizide |

| | |
|---|---|
| | empagliflozin |
| | sitagliptin |
| | dulaglutide |
| | empagliflozin |
| | semaglutide |
| | liraglutide |
| | empagliflozin |
| | sitagliptin |
| | glimepiride |
| | glyburide |
| | linagliptin |
| | canagliflozin |
| | alogliptin |
| | dapagliflozin |
| | exenatide |
| | repaglinide |
| | saxagliptin |
| | rosiglitazone |
| | acarbose |
| | ertugliflozin |
| | rosiglitazone |
| | nateglinide |
| | chlorpropamide |
| | albiglutide |
| | troglitazone |
| **Hypertension drugs** | lisinopril |
| | fosinopril |
| | benazepril |
| | captopril |
| | moexipril |
| | perindopril |
| | quinapril |
| | ramipril |
| | trandolapril |

| | |
|---|---|
| | enalapril |
| | hydrochlorothiazide |
| | furosemide |
| | indapamide |
| | chlorthalidone |
| | spironolactone |
| | trichlormethiazide |
| | chlorothiazide |
| | bumetanide |
| | bendroflumethiazide |
| | torsemide |
| | metolazone |
| | triamterene |
| | eplerenone |
| | amiloride |
| | hydralazine |
| | minoxidil |
| | aliskiren |
| | metoprolol |
| | labetalol |
| | atenolol |
| | nadolol |
| | nebivolol |
| | carvedilol |
| | propranolol |
| | esmolol |
| | bisoprolol |
| | acebutolol |
| | betaxolol |
| | timolol |
| | sotalol |
| | celiprolol |
| | pindolol |
| | penbutolol |
| | carteolol |

| | |
|---|---|
| | losartan |
| | valsartan |
| | olmesartan |
| | irbesartan |
| | candesartan |
| | telmisartan |
| | diltiazem |
| | amlodipine |
| | verapamil |
| | nifedipine |
| | nisoldipine |
| | diltiazem |
| | Felodipine |
| | isradipine |
| | felodipine |
| | nicardipine |
| | nimodipine |
| | clonidine |
| | methyldopa |
| | prazosin |
| | guanabenz |
| | guanfacine |
| | phentolamine |
| | prazosin |
| | doxazosin |
